# Diabetes related emergency department visits among adults during the COVID-19 pandemic – an analysis of data from the New York City syndromic surveillance system

**DOI:** 10.1101/2023.12.20.23300289

**Authors:** Anna Zhilkova, Ramona Lall, Robert Mathes, Shadi Chamany, Donald Olson

## Abstract

**Objective:** New York City was an early epicenter of the COVID-19 pandemic. We aim to describe population level epidemiological trends in diabetes related emergency department (ED) visits among adults in New York City, for the period prior to and encompassing the first four waves of the pandemic.

**Research Design and Methods:** We used data from the New York City ED syndromic surveillance system during December 30, 2018 through May 21, 2022. This system captures all visits from EDs in the city in near-real time. We matched these visits to laboratory confirmed COVID-19 positivity data beginning with February 15, 2020.

**Results:** Compared to pre-pandemic baseline levels, diabetes related ED visits noticeably increased during the first wave in spring 2020, though this did not necessarily translate to net increases overall during that period. Visits for diabetic ketoacidosis, particularly among adults with type 2 diabetes, sharply increased before returning to pre-pandemic levels, most notably during wave 1 and wave 4 in winter 2021-2022. Trajectories of diabetes-related ED visits differed by diabetes type, age, and sex. Some ED visit trends did not return to pre-pandemic baseline levels.

**Conclusions:** The COVID-19 pandemic, especially the first wave in the spring of 2020, coincided with a dramatic shift in diabetes related ED utilization in New York City. Our findings highlight the importance of on-going surveillance of health care utilization for chronic diseases during population-level emergencies like pandemics. A robust syndromic surveillance system that includes infectious and non-infectious syndromes is useful to better prepare, mitigate, and respond to population-level events.

**Article Highlights:**

- Diabetes related emergency department (ED) visits in New York City increased dramatically with the emergence of the COVID-19 pandemic in spring 2020.
- The trajectory of diabetes-related ED visits differed by diabetes type, age, sex, and pandemic wave.
- The diabetes complication of diabetic ketoacidosis among adults with type 2 diabetes showed sharp increases in the first and fourth waves of the pandemic, respectively its initial emergence in spring 2020 and the Omicron variant in winter 2021-2022.
- Our findings highlight the importance of on-going surveillance of health care utilization for chronic diseases during population-level emergencies like pandemics.

**Summary:** Data from NYC’s syndromic surveillance system showed major increases in #type2diabetes complications (e.g. diabetic ketoacidosis) during #COVID-19 waves 1 and 4 (Omicron) – this tool may be useful for population-level monitoring of chronic disease complications during emergencies

---

New York City was an early epicenter of the COVID-19 pandemic. During the initial stage, in the spring of 2020, the city experienced a dramatic rise in infections, hospitalizations, and deaths due to COVID-19, and greater excess mortality than elsewhere in the US (1–3). Emergency departments (EDs) in the city saw a drop in overall visits that coincided with lockdowns beginning in mid-March (4), similar to declines in ED utilization documented across the US and worldwide, though with a magnitude varying by severity, type of visit, reasons for visit, and other characteristics (5–8). Later reports demonstrated that some of the shifts persisted – there is much that remains to be understood about these longer-term trends in ED utilization (9).

Emergency rooms serve as gateways to other hospital services such as intensive care units, acute care hospitalization, and, beyond, rehabilitation and long-term care. During the pandemic, emergency rooms had to adapt to unique challenges. Characterizing ED use remains an important component of understanding the impact of the pandemic, as well as for informing preparedness efforts for future public health emergencies.

Particular attention has been devoted to changes in health care needs and experiences of people with diabetes during the pandemic. Population-based studies have found that diabetes related emergencies, most notably related to complications such as diabetic ketoacidosis, declined among people with type 1 diabetes and remained below pre-pandemic levels. Diabetic ketoacidosis among people with type 2 diabetes, however, increased and remained elevated (10,11). Some investigators have explained these trends by demonstrating that glycemic control has remained stable or improved among people with type 1 diabetes, while glycemic control among people with type 2 diabetes deteriorated (12,13). Others have pointed out that there is still much to learn about causal mechanisms behind shifts in diabetes emergencies, such as the role of a COVID-19 infection versus other contributing factors related to the pandemic such as delays in care or shifts in diabetes self-management (14,15).

In addition to the short-term effects of COVID-19, the emergence of long-term consequences of COVID-19 or ‘long COVID,’ including an increase in previously undiagnosed diabetes among adults may have implications for downstream increases in complications and health care utilization (16,17). The disparate impact of COVID-19 illness, complications, and mortality among Black, Latino, and other groups related to social, economic, and structural factors, has also been reported for diabetes related complications and shifts in health care (18–20).

There are several challenges with studying population-level trends of emergency health care utilization. First, while centralized systems of health care delivery, such as those found in the United Kingdom, allow for analysis of population-level analyses on different time scales, they are not available everywhere. Second, there is a need to understand both short- and long-term pandemic trends, particularly among populations most severely impacted. Widely used data sources such as hospital discharge data often fail to meet the need, as the lag in analysis often ranges from months to years.

Syndromic surveillance systems present an alternative for monitoring trends in chronic diseases such as diabetes. While the design of these systems have varied, in some places such as New York City, syndromic surveillance has enabled near real-time tracking of population-level ED use, providing critical information that may serve as an early warning. These systems have been used to monitor for early indicators of disease outbreaks, seasonal influenza, and understanding hospital utilization patterns during emergencies (21–23). With some exceptions, there has been less exploration and evaluation using syndromic surveillance to systematically track patterns of chronic disease-related ED use. In some of those instances, methodological approaches were often constrained by availability of diagnostic codes on the reported data and instead relied more on text-based reporting of syndromes (24–26). Over the past decade in New York City, integration of International Classification of Diseases, Tenth Revision, Clinical Modification (ICD-10) coding as well as improvements in hospital reporting have made the use of such tools for purposes like monitoring of chronic disease complications more tenable.

The aim of this study is to use population-level syndromic surveillance data to characterize changes in diabetes related ED visits among adults with type 1 and type 2 diabetes during the first four waves of the COVID-19 pandemic in New York City relative to pre-pandemic periods. Our results can be used to inform the next generation of syndromic surveillance systems that integrate infectious and non-infectious conditions. These syndromic surveillance systems may enable the near-real-time analysis of “syndemic” impacts and inform locally tailored, health equity interventions (27).

## Research Design and Methods

### Data

We used data from the New York City ED syndromic surveillance system. This system captures all visits from all 53 EDs citywide in near real time from electronic health records (23). Each patient record includes information such as date and time of visit, hospital facility, discharge diagnoses (available in about 90% of records) based on ICD-10 codes, discharge status, and patient demographics. For the present analysis, we include visits from December 30, 2018 through May 21, 2022.

Syndromic surveillance ED data were matched to COVID-19 positive laboratory results within the DOHMH’s communicable disease reporting system from February 15, 2020 onward. From February 15 to April 2020, data were matched on patient’s date of birth, sex, and zip code (patient names were not available during this period.) Beginning in April 2020, data were matched on patient’s date of birth, first name, and last name.

### Identifying Diabetes Related Visits

We classified ED visits for adults ages 18 and over into type 1 diabetes and type 2 diabetes using ICD-10 diagnostic codes E10x and E11x, respectively. We utilized Clinical Classification Software Refined (CCSR) version 2021.1 default outpatient categories to group visits into diabetes with complications (such as diabetic ketoacidosis, foot ulcers, hyperglycemia, hyperosmolarity, and retinopathy) and diabetes without complications. These together are meant to capture all diabetes related visits (28). Since the principal diagnosis is not available in syndromic data, we relied on all listed codes in the discharge diagnosis field. This means inclusion of records where diabetes is the reason for a visit, as well as where diabetes may be a secondary diagnosis. We thus use the term ‘diabetes related visits’ throughout. In some cases, the sum of visits classified as diabetes with complications and diabetes without complications may slightly exceed the total, because some records (fewer than 10%) may contain diagnostic codes for both.

We also report on several specific types of visits that fall under the diabetes with complications classification. For type 1 diabetes, we identified diabetic ketoacidosis visits, and for type 2 diabetes, we identified diabetic ketoacidosis, hyperglycemic hyperosmolar syndrome, as well a measure that combines the two, which many studies report as hyperglycemic crisis. Lastly, we included ICD-9 diagnostic codes corresponding to the categories described earlier, because we found that some hospital records still reported them. A complete list of diagnostic codes and definitions may be found in Table A5 in the supplementary materials.

### Selection of Time Periods

The pre-pandemic and pandemic periods were defined as follows:

- Pre-pandemic baseline: Dec 29, 2019-Feb 29, 2020 (CDC weeks 1-9 of 2020)
- Wave 1 (early pandemic period): Mar 1-May 30, 2020 (CDC weeks 10-22 of 2020)
- Wave 2: Nov 1 2020-May 22 2021 (CDC weeks 45-53 of 2020 and weeks 1-20 of 2021)
- Wave 3 (Delta variant): Jul 4-Nov 27, 2021 (CDC weeks 27-47 of 2021)
- Wave 4 (Omicron variant): Nov 28, 2021-Jan 1, 2022 (CDC weeks 48-52 of 2021, and Jan 2-Feb 26, 2022, or weeks 1-8 of 2022)

We chose to define time periods described above for the following reasons. The first laboratory confirmed case of COVID-19 in New York City was diagnosed on Feb 29, 2020. Although the introduction of the virus into the city likely occurred weeks earlier, rapid acceleration in COVID-19 related hospital use and excess mortality was not seen until March, peaking in April, and having largely fallen by end of May (1,2,20). New York City experienced a longer but less severe second wave beginning in the later part of 2020. We chose a cut-point in early November because this period similarly coincided with increased COVID-19 related hospital use. Similar rationale was used to define subsequent COVID-19 waves for the purposes of examining trends in ED use. While data for 2019 and earlier years were available, we restricted our baseline comparison to the early part of 2020 because it provided more stable reporting and consistent use of diabetes diagnostic codes by hospitals.

### Measures and Outcomes

For each visit type, we present a time series illustrating the number of weekly visits from January 2019 through May 2022. Additionally, for each period, we report mean weekly visits, percent of visits that are COVID-19 positive, percent of visits resulting in admissions, crude rate out of all ED visits, and total number of visits. Lastly, we disaggregate our findings by age and sex. We do not present data by race/ethnicity due to changes in reporting of this variable during the study period.

## Results

### Adults with Type 1 Diabetes

There was a decline in overall type 1 diabetes related visits among adults over time. The mean weekly number of visits was 55.2 (CI 95%, 49.5-60.9) during the pre-pandemic period in winter 2020, 49.4 (CI 95%, 40.5-58.3) during wave 1, 42.5 (95% CI, 39.7-45.3) during wave 2, 43.4 (CI 95% 40.2-46.7) during wave 3, and 42.1 (38.7-45.5) during wave 4 (Table 1.)

**Table 1:** Type 1 diabetes-related emergency department visits among adults ages 18 and older in New York City, December 29, 2019-February 26, 2022.

|  | **Pre-** | **pandemic** | **Wave 1** |  | **Wave 2** |  | **Wave 3** |  | **Wave 4** |  |
| --- | --- | --- | --- | --- | --- | --- | --- | --- | --- | --- |
|  | Dec 29, 2019-Feb 29, 2020 |  | Mar 1, 2020-May 30, 2020 |  | Nov 1, 2020-May 22, 2021 |  | Jul 4, 2021-Nov 27, 2021 |  | Nov 28, 2021-Feb 26, 2022 |  |
| **<u>Weekly means by patient characteristics</u>** |  |  |  |  |  |  |  |  |  |  |
| Ages 18-39 | 30.1 | (25.5-34.7) | 24.6 | (20.4-28.8) | 24.7 | (22.6-26.8) | 26 | (23.1-29) | 26.5 | (23.7-29.4) |
| Ages 40-64 | 17.3 | (14.7-20) | 17.5 | (13.3-21.7) | 12.9 | (11.5-14.3) | 13.4 | (11.8-15.1) | 12.2 | (10.7-13.8) |
| Ages 65+ | 7.8 | (5.8-9.8) | 7.3 | (4.5-10.1) | 4.9 | (4.3-5.4) | 4 | (2.9-5) | 3.3 | (2.4-4.2) |
| Female | 26.9 | (24.1-29.7) | 21.5 | (17.6-25.4) | 19.3 | (17.8-20.8) | 20.9 | (18.5-23.3) | 19.3 | (17.2-21.5) |
| Male | 28.3 | (24-32.6) | 27.9 | (22.7-33.2) | 23.1 | (21.3-25) | 22.5 | (20.7-24.4) | 22.8 | (19.3-26.2) |
| **<u>ED visit characteristics</u>** |  |  |  |  |  |  |  |  |  |  |
| **Diabetes without complications** |  |  |  |  |  |  |  |  |  |  |
| Weekly mean | 8.3 | (6.5-10.1) | 8.5 | (6.4-10.5) | 10.6 | (9.2-11.9) | 8.6 | (7.4-9.9) | 8.1 | (6.7-9.5) |
| % COVID-19 positive | 0 | (0-0) | 16.4 | (9.5-23.3) | 7.2 | (4.3-10.1) | 3.3 | (0.7-5.9) | 15.2 | (8.4-22.1) |
| Crude rate per 100K ED visits | 12.5 | (9.7-15.3) | 19.1 | (15.5-22.7) | 19.3 | (17.1-21.5) | 14.5 | (12.4-16.6) | 13.9 | (11.2-16.6) |
| **Diabetes with complications** |  |  |  |  |  |  |  |  |  |  |
| Weekly mean | 47.8 | (42.1-53.5) | 42.8 | (34.9-50.6) | 32.8 | (30.4-35.2) | 35.8 | (32.7-38.9) | 35.1 | (31.9-38.3) |
| % COVID-19 positive | 0 | (0-0) | 21.4 | (18-24.8) | 5.2 | (3.7-6.6) | 3.9 | (2.5-5.2) | 15.4 | (12-18.7) |
| Crude rate per 100K ED visits | 71.9 | (65.1-78.7) | 96.5 | (88.5-104.5) | 59.9 | (56.1-63.7) | 60.1 | (55.8-64.4) | 60.2 | (54.7-65.7) |
| **DKA** |  |  |  |  |  |  |  |  |  |  |
| Weekly mean | 26 | (22-30) | 24.7 | (19.4-30) | 15.8 | (14.2-17.5) | 17.8 | (15.8-19.9) | 20.8 | (18.5-23.2) |
| % COVID-19 positive | 0 | (0-0) | 23.1 | (18.4-27.7) | 7 | (4.6-9.3) | 4.5 | (2.4-6.7) | 17 | (12.5-21.4) |
| Crude rate per 100K ED visits | 39.1 | (34.1-44.1) | 55.7 | (49.6-61.8) | 28.9 | (26.3-31.5) | 29.9 | (26.9-32.9) | 35.8 | (31.5-40.1) |
| **Totals** |  |  |  |  |  |  |  |  |  |  |
| Weekly mean | 55.2 | (49.5-60.9) | 49.4 | (40.5-58.3) | 42.5 | (39.7-45.3) | 43.4 | (40.2-46.7) | 42.1 | (38.7-45.5) |
| % COVID-19 positive | 0 | (0-0) | 20.4 | (17.3-23.5) | 5.7 | (4.4-7) | 3.8 | (2.6-5.1) | 15.4 | (12.3-18.4) |
| Crude rate per 100K ED visits | 83.1 | (75.8-90.4) | 111.5 | (102.9-120.1) | 77.6 | (73.3-81.9) | 72.9 | (68.2-77.6) | 72.2 | (66.1-78.3) |
| % admitted | 63.4 | (59.1-67.6) | 60.6 | (56.8-64.4) | 47 | (44.2-49.8) | 45.9 | (42.7-49.2) | 51 | (46.8-55.2) |
| N | 497 |  | 642 |  | 1232 |  | 912 |  | 547 |  |

Abbreviations: DKA = diabetic ketoacidosis; ED = emergency department.
COVID-19 positivity was ascertained through linkage to laboratory confirmed data.
Crude rates were calculated based on total adult ED visits for each period.
Weekly means, rates, and percentages are presented to three or fewer significant digits.
95% confidence intervals in parentheses.

Trends in overall type 1 diabetes related visits during wave 1 of the pandemic mask nuanced changes within that time frame. For every visit type – without complications, with complications, and diabetic ketoacidosis – there were sharp increases followed by declines during some weeks of wave 1 (Figure 1.) However, despite these shifts during some weeks, there were no net increases during wave 1, as the mean weekly visits were at similar levels or somewhat below pre-pandemic baseline for every visit type (Table 1.) After wave 1, visits for diabetes without complications remained at similar levels for the remainder of the study period, while visits for diabetes with complications and visits for diabetic ketoacidosis declined.

**Figure 1:**
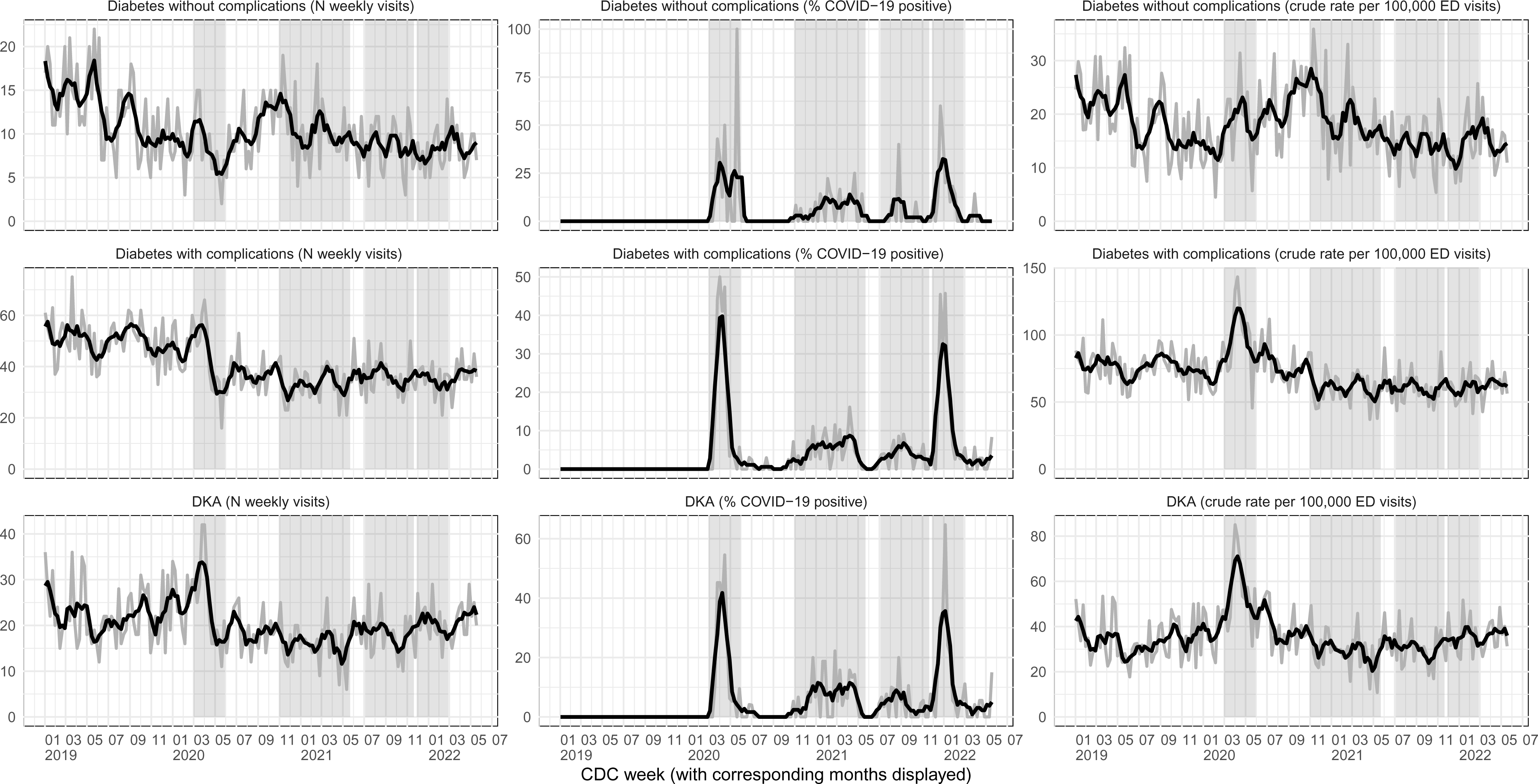
Type 1 diabetes-related emergency department visits among adults ages 18 and older in New York City, 2019−2022. Abbreviations: DKA = diabetic ketoacidosis; ED = emergency department. Gray line = weekly value; black line = four week average; shaded regions = COVID−19 waves of ED visits in New York City (wave 1: March 1, 2020−May 30, 2020, wave 2: November 1, 2020−May 22, 2021, wave 3: July 4, 2021− November 27, 2021, wave 4: November 28, 2021−February 26, 2022.) Diabetes−related ED visits were identified based on any listed diagnostic codes in the discharge diagnosis field (available in about 90% of records) of syndromic surveillance data from all New York City EDs. Diabetes without complications and diabetes with complications groupings are meant to capture all diabetes related visits and are based on Clinical Classification Software Revised (CCSR) for ICD−10 diagnoses, version 2021.1. DKA is a subset of diabetes with complications. COVID−19 positivity was ascertained through linkage to laboratory confirmed data. Denominator for crude rates = total weekly adult ED visits.

Every age group saw a decline in overall type 1 diabetes related visits over time compared to the pre-pandemic period, though the timing of these shifts varied. Women and men had similar levels of overall type 1 diabetes related visits during the pre-pandemic period. Visits among women declined during wave 1, while among men remained at similar levels to the pre-pandemic period. By the end of our study period, visits among both women and men were below pre-pandemic levels (Table 1.) For type 1 diabetic ketoacidosis, there were no increases in visits observed during wave 1 or subsequent periods among women or men, or among 18-39 and 65+ age groups, though there was an increase among the 40-64 age group (Table A1.)

In examining trends in COVID-19 infections, we found that percent of diabetes-related ED visits that were COVID-19 positive lined up with increases in COVID-19 hospital use during waves 1-4, as expected (Figure 1.) Increases in COVID-19 positivity did not appear to coincide with net increases in type 1 diabetes related visits (Table 1.) Additionally, we found an increase in the crude rate of type 1 diabetes with complications visits out of all ED visits among adults during wave 1 – even as the mean weekly visits remained at similar levels to the pre-pandemic period (Table 1.) This may be reflective of a larger drop in total ED visits (denominator) in New York City, even as people may have continued going to the ED for higher acuity events.

### Adults with Type 2 Diabetes

There was a decline in overall type 2 diabetes related visits among adults over time, similar to trends observed for overall type 1 diabetes related visits. The mean weekly number of visits was 996 (95% CI, 964-1030) during the pre-pandemic period in winter 2020, 961 (95% CI, 816-1110) during wave 1, 851 (95% CI, 835-866) during wave 2, 737 (CI 95% 707-766) during wave 3, and 703 (CI 95% 666-741) during wave 4 (Table 2.)

**Table 2:** Type 2 diabetes-related emergency department visits among adults ages 18 and older in New York City, December 29, 2019-February 26, 2022.

|  | **Pre-** | **pandemic** | **Wave 1** |  | **Wave 2** |  | **Wave 3** |  | **Wave 4** |  |
| --- | --- | --- | --- | --- | --- | --- | --- | --- | --- | --- |
|  | Dec 29, 2019-Feb 29, 2020 |  | Mar 1, 2020-May 30, 2020 |  | Nov 1, 2020-May 22, 2021 |  | Jul 4, 2021-Nov 27, 2021 |  | Nov 28, 2021-Feb 26, 2022 |  |
| **<u>Weekly means by patient characteristics</u>** |  |  |  |  |  |  |  |  |  |  |
| Ages 18-39 | 105 | (96.3-113) | 86.9 | (75.0-98.8) | 98.1 | (93.5-103) | 94.9 | (89.6-100) | 88.7 | (83.8-93.6) |
| Ages 40-64 | 471 | (445-497) | 452 | (384-520) | 434 | (425-443) | 369 | (353-386) | 356 | (338-373) |
| Ages 65+ | 420 | (406-435) | 422 | (351-493) | 319 | (309-329) | 273 | (261-285) | 259 | (239-278) |
| Female | 495 | (475-515) | 432 | (366-497) | 394 | (384-403) | 341 | (327-356) | 322 | (299-345) |
| Male | 501 | (485-517) | 529 | (446-612) | 457 | (447-467) | 396 | (379-413) | 381 | (365-397) |
| **<u>ED visit characteristics</u>** |  |  |  |  |  |  |  |  |  |  |
| **<u>Diabetes without complications</u>** |  |  |  |  |  |  |  |  |  |  |
| Weekly mean | 500 | (479-522) | 500 | (420-580) | 471 | (463-479) | 390 | (365-414) | 365 | (340-391) |
| % COVID-19 positive | 0 | (0-0) | 23.4 | (22.4-24.4) | 8.5 | (8.1-9.0) | 3.6 | (3.2-4.0) | 15 | (14-16) |
| Crude rate per 100K ED visits | 753 | (731-775) | 1130 | (1100-1160) | 859 | (845-874) | 654 | (640-668) | 627 | (609-644) |
| **<u>Diabetes with complications</u>** |  |  |  |  |  |  |  |  |  |  |
| Weekly mean | 566 | (550-583) | 521 | (442-601) | 412 | (399-426) | 376 | (363-389) | 362 | (344-380) |
| % COVID-19 positive | 0 | (0-0) | 27.5 | (26.4-28.6) | 8.6 | (8.1-9.1) | 2.6 | (2.2-2.9) | 15.5 | (14.4-16.5) |
| Crude rate per 100K ED visits | 852 | (828-875) | 1180 | (1150-1200) | 753 | (740-767) | 632 | (618-646) | 621 | (603-639) |
| **<u>DKA</u>** |  |  |  |  |  |  |  |  |  |  |
| Weekly mean | 69.9 | (62.1-77.6) | 115 | (81.5-148) | 69.9 | (66.2-73.7) | 69.1 | (65.3-72.9) | 83.8 | (74.3-93.4) |
| % COVID-19 positive | 0 | (0-0) | 39.2 | (36.7-41.7) | 14.3 | (12.8-15.9) | 5.5 | (4.3-6.7) | 22.9 | (20.4-25.4) |
| Crude rate per 100K ED visits | 105 | (96.9-113) | 259 | (246-273) | 128 | (122-133) | 116 | (110-122) | 144 | (135-152) |
| **<u>HHS</u>** |  |  |  |  |  |  |  |  |  |  |
| Weekly mean | 10.4 | (8.4-12.5) | 15.1 | (10.9-19.2) | 13.6 | (12.1-15.1) | 11.9 | (10.7-13.1) | 15.9 | (13.7-18.2) |
| % COVID-19 positive | 0 | (0-0) | 32.7 | (26.1-39.2) | 11.4 | (8.3-14.6) | 1.2 | (0-2.5) | 20.3 | (14.8-25.8) |
| Crude rate per 100K ED visits | 15.7 | (12.5-18.9) | 34 | (29.2-38.8) | 24.8 | (22.4-27.2) | 20 | (17.5-22.5) | 27.3 | (23.6-31) |
| **<u>Hyperglycemic crisis (DKA or HHS)</u>** |  |  |  |  |  |  |  |  |  |  |
| Weekly mean | 79.7 | (72.5-86.8) | 128 | (91.4-164) | 81.5 | (77.7-85.3) | 79.8 | (75.9-83.6) | 97.5 | (87.1-108) |
| % COVID-19 positive | 0 | (0-0) | 38.5 | (36.1-40.8) | 13.8 | (12.4-15.1) | 5 | (3.9-6) | 22.4 | (20.1-24.7) |
| Crude rate per 100K ED visits | 120 | (111-129) | 288 | (274-302) | 149 | (143-155) | 134 | (128-140) | 167 | (158-177) |
| **<u>Totals</u>** |  |  |  |  |  |  |  |  |  |  |
| Weekly mean | 996 | (964-1030) | 961 | (816-1110) | 851 | (835-866) | 737 | (707-767) | 703 | (666-741) |
| % COVID-19 positive | 0 | (0-0) | 24.8 | (24.1-25.6) | 8.5 | (8.1-8.8) | 3.1 | (2.8-3.4) | 15.1 | (14.4-15.9) |
| Crude rate per 100K ED visits | 1500 | (1470-1530) | 2170 | (2130-2210) | 1550 | (1530-1570) | 1240 | (1220-1260) | 1210 | (1180-1230) |
| % admitted | 43.3 | (42.3-44.3) | 40.4 | (39.6-41.3) | 32.3 | (31.7-32.9) | 30.7 | (30-31.4) | 31.6 | (30.6-32.5) |
| N | 8966 |  | 12490 |  | 24672 |  | 15478 |  | 9141 |  |

Abbreviations: DKA = diabetic ketoacidosis; HHS = hyperosmolar hyperglycemic syndrome; ED = emergency department.
COVID-19 positivity was ascertained through linkage to laboratory confirmed data.
Crude rates were calculated based on total adult ED visits for each period.
Weekly means, rates, and percentages are presented to three or fewer significant digits.
95% confidence intervals in parentheses.

Trends in overall type 2 diabetes related visits also mask important differences in changes that occurred over time. For every visit type – diabetes without complications, diabetes with complications, diabetic ketoacidosis, and hyperglycemic hyperosmolar syndrome – there were some sharp increases in visits followed by declines within wave 1 (Figure 2.) Despite notable shifts during some weeks of wave 1, there were no net increases for visits related to diabetes without complications, or diabetes with complications: the mean weekly visits remained comparable or somewhat below pre-pandemic levels (Table 2.)

**Figure 2:**
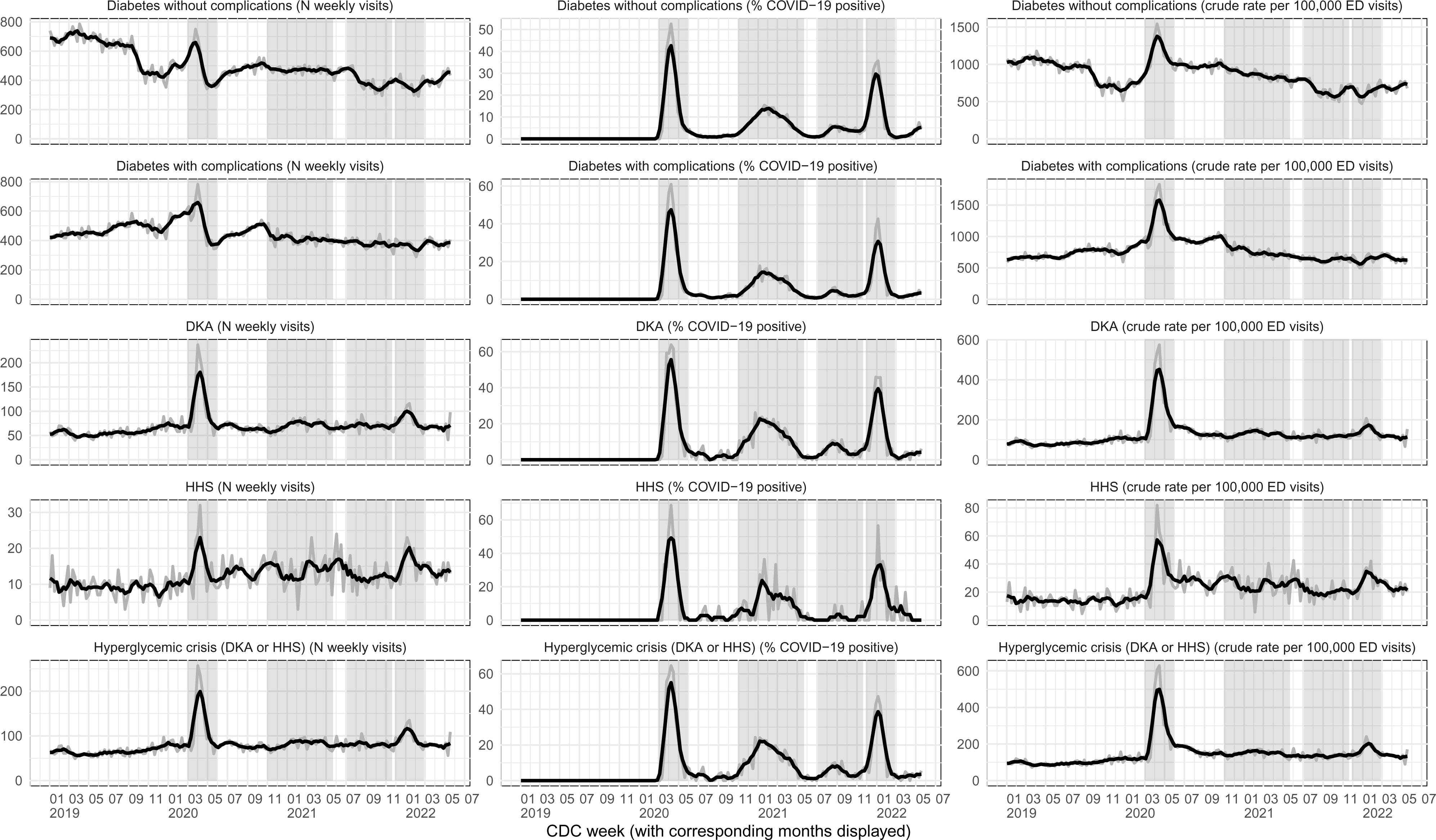
Type 2 diabetes-related emergency department visits among adults ages 18 and older in New York City, 2019−2022. Abbreviations: DKA = diabetic ketoacidosis; HHS = hyperosmolar hyperglycemic syndrome; ED = emergency department. Gray line = weekly value; black line = four week average; shaded regions = COVID−19 waves of ED visits in New York City (wave 1: March 1, 2020−May 30, 2020, wave 2: November 1, 2020−May 22, 2021, wave 3: July 4, 2021− November 27, 2021, wave 4: November 28, 2021−February 26, 2022.) Diabetes−related ED visits were identified based on any listed diagnostic codes in the discharge diagnosis field (available in about 90% of records) of syndromic surveillance data from all New York City EDs. Diabetes without complications and diabetes with complications groupings are meant to capture all diabetes related visits and are based on Clinical Classification Software Revised (CCSR) for ICD−10 diagnoses, version 2021.1. DKA, HHS, hyperglycemic crisis = subsets of diabetes with complications. COVID−19 positivity was ascertained through linkage to laboratory confirmed data. Denominator for crude rates = total weekly adult ED visits.

A stark exception to these trends is type 2 diabetic ketoacidosis, which makes up a small proportion of diabetes with complications visits (about 10% in our data,) but for which visits increased during wave 1 by over 60%, before returning to pre-pandemic baseline levels. The mean weekly number of visits for type 2 diabetic ketoacidosis was 69.9 (95% CI, 62.1-77.6) during the pre-pandemic period, 115 (95% CI, 81.5-148) during wave 1, 69.9 (95% CI, 66.2-73.7) during wave 2, 69.1 (CI 95% 65.3-72.9) during wave 3, and 83.8 (CI 95% 74.3-93.4) during wave 4 (Table 2.) Similar trends were also observed for hyperglycemic hyperosmolar syndrome for wave 1.

Several observations are noteworthy about trends for type 2 diabetic ketoacidosis. First, there are some departures from trends observed for type 1 diabetes. During one week of wave 1, visits related to type 2 diabetic ketoacidosis tripled to over 200 per week, before returning to near pre-pandemic levels by the conclusion of wave 1 (Figure 2.) Second, the prominent increases observed during wave 1 are driven by 40-64 and 65+ age groups, while no change was found for the 18-39 group. Among the 40-64 age group, visits increased by about 80%, from a pre-pandemic weekly mean of 29.6 (95% CI 24.8-34.4) to 53.8 (95% CI 36.6-71.0) during wave 1, while among the 65+ age group, visits increased by over 130%, from a pre-pandemic weekly mean of 15.4 (95% CI 13.8-17.0) to 36.6 (95% CI 21.2-52.0) (Table A2.) During wave 4, visits were also unchanged among the 18-39 group, while both the 40-64 and 65+ age groups experienced increases. Further analysis (not shown) revealed that excluding COVID-19 positive visits (39.2% of type 2 diabetic ketoacidosis visits during wave 1,) eliminated much of the dramatic increase observed in type 2 diabetic ketoacidosis during wave 1.

After wave 1, type 2 diabetes related visits with and without complications declined, and remained below pre-pandemic levels through the end of the study period in early 2022. Some types of visits, such as for diabetic ketoacidosis and hyperosmolar hyperglycemic syndrome, increased during periods of high COVID-19 activity. Since visits related to diabetic ketoacidosis and hyperosmolar hyperglycemic syndrome make up a relatively small proportion of type 2 diabetes complications, those increases are less likely to influence overall trends and may be more challenging to detect with more broadly aggregate data.

Overall type 2 diabetes related visits appeared to decline for adults of all ages, though the timing varied by age group, with the biggest changes observed for the 65+ group following wave 1 (Table 2.) Visit levels were nearly the same for women and men during the pre-pandemic period, however, visits among women declined beginning with wave 1, while visits among men increased. After wave 1, visits among both women and men declined, though visits among men remained at higher levels compared to women. Trends by race/ethnicity were not examined due to missing data and variation in reporting of race and ethnicity by hospitals over time.

Patterns of COVID-19 disease and type 2 diabetes related visits with and without complications shared similarities with those of type 1 diabetes related visits, in that we found increases in the proportion of visits with COVID-19 during waves 1-4 as expected, with the biggest increases in positivity occurring during wave 1 and wave 4. These increases in COVID-19 coincided with notable increases in some type 2 diabetes related complications.

## Conclusions

The COVID-19 pandemic resulted in substantial shifts in diabetes related ED use among adults in New York City. We found that both type 1 and type 2 diabetes related visits declined over the course of the study period beginning with the pre-pandemic period in early 2020 through the conclusion of wave 4 in the early part of 2022. An important exception to these overall trends was that visits related to some type 2 diabetes complications such as diabetic ketoacidosis and hyperosmolar hyperglycemic syndrome – both of which increased substantially during waves 1 and 4 – were largely among COVID-19 positive patients. Trends of diabetic ketoacidosis in New York City are broadly consistent with those reported in other population-level studies, although our findings suggest there was a return closer to pre-pandemic levels during periods of reduced COVID-19 activity (9,10).

Our findings further corroborate that utilization by patients with type 1 diabetes differed from those with type 2 diabetes in other important ways. For example, we did not observe net increases in diabetic ketoacidosis among patients with type 1 diabetes during any COVID-19 wave during our study period, although there were substantial increases among patients with type 2 diabetes during periods of high infection rates. Furthermore, these differences may not be fully explained by age, as we did not observe increases among patients 65 and older with type 1 diabetes, while there were substantial increases among this age group among patients with type 2 diabetes. Lastly, our findings about overall declines in adult diabetes related ED visits are consistent with trends that have been reported about health care utilization for other chronic diseases (29).

The pandemic had a devastating impact on overall mortality, both directly through COVID-19 infections and through indirect effects, such as consequences of a stressed healthcare system, and changes in health care utilization (30). Given that different populations were impacted during different stages of the pandemic, and that severity of cases combined with restrictions also varied with time, careful examination is warranted to tease out causal effects of these factors. Some findings, such as a notable increase in type 2 diabetic ketoacidosis and hyperosmolar hyperglycemic syndrome during wave 4 (first omicron variant) – in absence of lockdowns – suggest a direct role of COVID-19 infections, as opposed to indirect effects such as interruptions in availability of services or factors such as more gradual lifestyle changes, though it is possible that other barriers may have contributed to delays in routine care seeking. Other trends, such as declines in overall type 1 and 2 diabetes visits that persisted two years into the pandemic, raise several possibilities. One possibility is mortality displacement – whereby patients with uncontrolled diabetes and other comorbidities had higher mortality rates during earlier pandemic periods, while those who survived experienced fewer diabetes related emergencies. Additional contributing factors may include shifts in the way people access health care, such as increased adoption and use of telehealth services, or improvements in glucose control observed among people with type 1 diabetes (11, 13).

There are several limitations of our analysis. First, we did not have information about the principal diagnosis of each ED visit, and instead relied on all reported codes listed under the diagnosis field. Thus, we may be overestimating the total number of ED visits due to diabetes, by capturing some visits among individuals with diabetes as a comorbidity, but who presented to the emergency room for other reasons.

Second, owing to concerns over reporting of some diabetes diagnostic codes in 2019, we relied on January and February of 2020 as our pre-pandemic baseline. A limitation may be that we are not adequately capturing seasonality, which has been reported in both diabetes mortality (30) and in the incidence of new onset type 2 diabetes among adults (31) Doro 2006.) To the extent that seasonality exists not just in diabetes mortality and incidence, but also in ED visits, it is possible that the early 2020 pre-pandemic period may ‘overestimate’ visit counts relative to other months.

Third, due to concerns about missing data and variation in reporting of race and ethnicity by hospitals over time, we are not able to quantify the extent of inequities by race and ethnicity that have been documented elsewhere (32). These data are essential in addressing syndemic effects and making targeted health equity investments in response to pandemics and other population-level events. The next generation of syndromic surveillance systems will need to improve demographic data collection and quality. The NYC Health Department is actively working with its partners to improve race and ethnicity data collection by reporting hospitals. As health systems express increasing commitments to health equity, one opportunity for improvement may be more accurate and systematic reporting of patient characteristics such as race/ethnicity (33).

Fourth, while we discuss trends among patients with type 1 and type 2 diabetes with respect to complications such as diabetic ketoacidosis, we are not able to explore potential contributing factors beyond characteristics such as age and COVID-19 positivity. Fifth, changes in population dynamics – as much as 5% of the total population, disproportionately from wealthier neighborhoods, left New York City in the early stages of the pandemic, combined with excess mortality among older New Yorkers during the same period – may impact some of the trends we are observing (34, 35). Lastly, though we offer some potential explanations for epidemiological trends presented here, we would caution against making causal claims without further inquiry into patient- and structural-level factors.

Syndromic surveillance systems present a valuable opportunity for timely monitoring of population-level trends in diabetes and other chronic disease complications. Among the leading causes of premature mortality before the pandemic, diabetes has been a major contributor to severe outcomes from SARS-CoV-2 infection, and the pandemic has had a catastrophic impact on persons living with diabetes (36). Monitoring changes related to diabetes and other chronic diseases will gain in importance in years to come, as there is growing evidence suggesting a heightened risk of developing diabetes following SARS-CoV-2 infection (37). Additionally, as some communities, including Black and Latino populations, were disproportionately impacted during the pandemic, using novel data systems to track health status more quickly and completely, with greater granularity, including geographic variation, may shed light on long-term shifts in health disparities and inform public health practice (38). The next generation of syndromic surveillance systems, managed by local and state health departments, with higher quality data, integration across the infectious and non-infectious syndrome continuum, and with highly accurate demographic information, will improve our capacity to respond to pandemics and their related syndemic factors (39).

## Data Availability

All data produced in the present study are available upon reasonable request to the authors through signed Data Use Agreement with the New York City Department of Health and Mental Hygiene, as required by Department of Health and Mental Hygiene guidelines.

## Supplemental Materials

Supplemental Table A1: Patient characteristics of type 1 diabetic ketoacidosis-related emergency department visits among adults ages 18 and older in New York City, December 29, 2019-February 26, 2022.

Supplemental Table A2: Patient characteristics of type 2 diabetic ketoacidosis-related emergency department visits among adults ages 18 and older in New York City, December 29, 2019-February 26, 2022.

Supplemental Table A3: Patient characteristics of type 2 hyperosmolar hyperglycemic syndrome-related emergency department visits among adults ages 18 and older in New York City, December 29, 2019-February 26, 2022.

Supplemental Table A4: Patient characteristics of type 2 hyperglycemic crisis (diabetic ketoacidosis or hyperosmolar hyperglycemic syndrome) -related emergency department visits among adults ages 18 and older in New York City, December 29, 2019-February 26, 2022.

Supplemental Table A5: List of diagnostic codes for classifying diabetes-related emergency department visits.

## Notes

### Competing Interest Statement

The authors have declared no competing interest.

### Funding Statement

This study did not receive any funding.

### Author Declarations

This study was classified as public health surveillance and exempt from ethical review and informed consent by the Institutional Review Board of the New York City Department of Health and Mental Hygiene.

