## Supplemental Table A1 for "Diabetes related emergency department visits among adults during the COVID-19 pandemic – an analysis of data from the New York City syndromic surveillance system"

Supplementary Table A1: Patient characteristics of type 1 diabetic ketoacidosis-related emergency department visits among adults ages 18 and older in New York City, December 29, 2019-February 26, 2022

|  | **Pre-** | **pandemic** | **Wave 1** |  | **Wave 2** |  | **Wave 3** |  | **Wave 4** |  |
| --- | --- | --- | --- | --- | --- | --- | --- | --- | --- | --- |
| **TYPE 1 DKA** | Dec 29, 2019-Feb 29, 2020 | | Mar 1, 2020-May 30, 2020 | | Nov 1, 2020-May 22, 2021 | | Jul 4, 2021-Nov 27, 2021 | | Nov 28, 2021-Feb 26, 2022 | |
| **Weekly means by patient characteristics** |  |  |  |  |  |  |  |  |  |  |
| Ages 18-39 | 17.1 | (13.5-20.7) | 13.1 | (10.2-15.9) | 9.4 | (8.2-10.5) | 11.6 | (9.6-13.5) | 13.8 | (11.5-16) |
| Ages 40-64 | 6.4 | (4.3-8.6) | 9.1 | (6.8-11.4) | 5 | (4.2-5.8) | 5.2 | (4.3-6.1) | 5.6 | (4.7-6.5) |
| Ages 65+ | 2.4 | (1.2-3.7) | 2.5 | (1.4-3.7) | 1.4 | (1.1-1.8) | 1 | (0.4-1.7) | 1.5 | (1-1.9) |
| Female | 11.4 | (8.7-14.1) | 10 | (7.6-12.4) | 7.5 | (6.6-8.4) | 9.1 | (7.6-10.7) | 9.8 | (8-11.5) |
| Male | 14.6 | (12.1-17) | 14.7 | (11.5-17.9) | 8.3 | (7.1-9.6) | 8.7 | (7.4-9.9) | 11.1 | (9-13.2) |

Abbreviations: ED = emergency department.

Diabetes-related ED visits were identified based on any listed diagnostic codes in the discharge diagnosis field (available in about 90% of records) of syndromic surveillance data from all New York City EDs.

Weekly means, rates, and percentages are presented to three or fewer significant digits.

95% confidence intervals in parentheses.
