## Supplemental Table A2 for "Diabetes related emergency department visits among adults during the COVID-19 pandemic – an analysis of data from the New York City syndromic surveillance system"

Supplementary Table A2: Patient characteristics of type 2 diabetic ketoacidosis-related emergency department visits among adults ages 18 and older in New York City, December 29, 2019-February 26, 2022

|  | **Pre-** | **-pandemic** | **Wave 1** |  | **Wave 2** |  | **Wave 3** |  | **Wave 4** |  |
| --- | --- | --- | --- | --- | --- | --- | --- | --- | --- | --- |
| **TYPE 2 DKA** | Dec 29, 2019-Feb 29, 2020 | | Mar 1, 2020-May 30, 2020 | | Nov 1, 2020-May 22, 2021 | | Jul 4, 2021-Nov 27, 2021 | | Nov 28, 2021-Feb 26, 2022 | |
| **Weekly means by patient characteristics** |  |  |  |  |  |  |  |  |  |  |
| Ages 18-39 | 24.9 | (20.5-29.3) | 24.5 | (21.9-27.2) | 19.3 | (17-21.7) | 22.1 | (19.4-24.9) | 22.4 | (20.2-24.6) |
| Ages 40-64 | 29.6 | (24.8-34.4) | 53.8 | (36.6-71) | 34 | (31.4-36.6) | 32.2 | (30.3-34.2) | 41.8 | (36.4-47.2) |
| Ages 65+ | 15.4 | (13.8-17) | 36.6 | (21.2-52) | 16.6 | (14.8-18.3) | 14.7 | (13.4-16) | 19.7 | (14.7-24.7) |
| Female | 28.3 | (23-33.7) | 42.5 | (30-55) | 29.3 | (26.9-31.7) | 31.3 | (28.6-34.1) | 37.6 | (32.2-43) |
| Male | 41.6 | (37.1-46) | 72.4 | (50.6-94.1) | 40.6 | (38.1-43.2) | 37.7 | (35.2-40.2) | 46.2 | (40.7-51.8) |

Abbreviations: ED = emergency department.

Diabetes-related ED visits were identified based on any listed diagnostic codes in the discharge diagnosis field (available in about 90% of records) of syndromic surveillance data from all New York City EDs.

Weekly means, rates, and percentages are presented to three or fewer significant digits.

95% confidence intervals in parentheses.
