## Supplemental Table A3 for "Diabetes related emergency department visits among adults during the COVID-19 pandemic – an analysis of data from the New York City syndromic surveillance system"

Supplementary Table A3: Patient characteristics of type 2 hyperosmolar hyperglycemic syndrome-related emergency department visits among adults ages 18 and older in New York City, December 29, 2019-February 26, 2022

|  | **Pre-** | **pandemic** | **Wave 1** |  | **Wave 2** |  | **Wave 3** |  | **Wave 4** |  |
| --- | --- | --- | --- | --- | --- | --- | --- | --- | --- | --- |
| **TYPE 2 HHS** | Dec 29, 2019-Feb 29, 2020 | | Mar 1, 2020-May 30, 2020 | | Nov 1, 2020-May 22, 2021 | | Jul 4, 2021-Nov 27, 2021 | | Nov 28, 2021-Feb 26, 2022 | |
| **Weekly means by patient characteristics** |  |  |  |  |  |  |  |  |  |  |
| Ages 18-39 | 1.2 | (0.6-1.9) | 1.2 | (0.3-2) | 1.2 | (0.8-1.7) | 1.4 | (0.8-2.1) | 1.8 | (1.2-2.3) |
| Ages 40-64 | 4.4 | (3-5.9) | 6.5 | (5-8) | 6.7 | (5.5-7.9) | 5.3 | (4.4-6.1) | 7.9 | (6.1-9.8) |
| Ages 65+ | 4.8 | (3-6.6) | 7.5 | (4.2-10.8) | 5.6 | (4.7-6.5) | 5.2 | (4.2-6.2) | 6.2 | (4.9-7.5) |
| Female | 3.9 | (2.5-5.3) | 6.8 | (4.8-8.9) | 6 | (5-7.1) | 5.2 | (4.5-6) | 8 | (7.2-8.8) |
| Male | 6.6 | (5.3-7.8) | 8.2 | (5.5-10.9) | 7.6 | (6.4-8.8) | 6.7 | (5.6-7.8) | 7.9 | (5.9-10) |

Abbreviations: ED = emergency department.

Diabetes-related ED visits were identified based on any listed diagnostic codes in the discharge diagnosis field (available in about 90% of records) of syndromic surveillance data from all New York City EDs.

Weekly means, rates, and percentages are presented to three or fewer significant digits.

95% confidence intervals in parentheses.
