## Supplemental Table A4 for "Diabetes related emergency department visits among adults during the COVID-19 pandemic – an analysis of data from the New York City syndromic surveillance system"

Supplementary Table A4: Patient characteristics of type 2 hyperglycemic crisis (diabetic ketoacidosis or hyperosmolar hyperglycemic syndrome) -related emergency department visits among adults ages 18 and older in New York City, December 29, 2019-February 26, 2022

|  | **Pre-** | **pandemic** | **Wave 1** |  | **Wave 2** |  | **Wave 3** |  | **Wave 4** |  |
| --- | --- | --- | --- | --- | --- | --- | --- | --- | --- | --- |
| **TYPE 2 DKAHHS** | Dec 29, 2019-Feb 29, 2020 | | Mar 1, 2020-May 30, 2020 | | Nov 1, 2020-May 22, 2021 | | Jul 4, 2021-Nov 27, 2021 | | Nov 28, 2021-Feb 26, 2022 | |
| **Weekly means by patient characteristics** |  |  |  |  |  |  |  |  |  |  |
| Ages 18-39 | 26 | (21.6-30.4) | 25.4 | (22.6-28.2) | 20.3 | (18-22.7) | 23.4 | (20.6-26.1) | 23.6 | (21.4-25.8) |
| Ages 40-64 | 33.6 | (29.3-37.8) | 59.1 | (41.6-76.6) | 39.7 | (37.1-42.3) | 36.9 | (34.9-38.8) | 48.5 | (42.6-54.3) |
| Ages 65+ | 20.1 | (18.6-21.7) | 43.2 | (25.1-61.2) | 21.4 | (19.5-23.4) | 19.5 | (17.9-21.1) | 25.5 | (20.4-30.5) |
| Female | 32.1 | (27.5-36.7) | 48.2 | (34.7-61.8) | 34.3 | (31.8-36.8) | 35.9 | (32.9-38.9) | 44.7 | (39.4-50) |
| Male | 47.6 | (43-52.1) | 79.4 | (56-102.8) | 47.1 | (44.4-49.9) | 43.8 | (41.6-46) | 52.8 | (46.2-59.5) |

Abbreviations: ED = emergency department.

Diabetes-related ED visits were identified based on any listed diagnostic codes in the discharge diagnosis field (available in about 90% of records) of syndromic surveillance data from all New York City EDs.

Weekly means, rates, and percentages are presented to three or fewer significant digits.

95% confidence intervals in parentheses.
