## Supplemental Table A5 for "Diabetes related emergency department visits among adults during the COVID-19 pandemic – an analysis of data from the New York City syndromic surveillance system"

Supplementary Table A5: List of diagnostic codes for classifying diabetes-related emergency department visits

|  |  | **ICD-10 codes beginning with:** | **ICD-9 codes beginning with:** |
| --- | --- | --- | --- |
| **Type 1 diabetes** | Diabetes without complications | Codes beginning with E10 under “END002” default outpatient category of Clinical Classification Software Revised (CCSR) version 2021.1 | 250.01 |
|  | Diabetes with complications | Codes beginning with E10 under “END003” default outpatient category of Clinical Classification Software Revised (CCSR) version 2021.1 | 250.03, 250.11, 250.13, 250.21, 250.23, 250.31, 250.33, 250.41, 250.43, 250.51, 250.53, 250.61, 250.63, 250.71, 250.73, 250.81, 250.83, 250.91, 250.93 |
|  | DKA | E10.1 | 250.11, 250.13 |
| **Type 2 diabetes** | Diabetes without complications | Codes beginning with E11 under “END002” default outpatient category of Clinical Classification Software Revised (CCSR) version 2021.1 | 250.00 |
|  | Diabetes with complications | Codes beginning with E11 under + “END003” default outpatient category of Clinical Classification Software Revised (CCSR) version 2021.1  E13.1 (DKA) | 250.02, 250.10, 250.12, 250.20, 250.22, 250.30, 250.32, 250.40, 250.42, 250.50, 250.52, 250.60, 250.62, 250.70, 250.72, 250.80, 250.82, 250.90, 250.92 |
|  | DKA | E11.1, E13.1, | 250.10, 250.12 |
|  | HHS | E11.0 | 250.02 |

Abbreviations: DKA = diabetic ketoacidosis; HHS = hyperosmolar hyperglycemic syndrome; ED = emergency department.

For type 2 DKA, code E13.1 (“other specified diabetes with ketoacidosis”) was included because the initial release of ICD-10 did not contain a specific code to identify type 2 DKA, and E13.1 was utilized instead until approximately 2018 (Carrier 2018, Dugan and Shubrook 2017.) Additionally, hospital adoption and reporting may not have been immediate, as some hospital records in our data contained ICD-9 codes in 2019 and after.
